# Prevalence of short interpregnancy interval and its associated factors among pregnant women in Debre Berhan town, Ethiopia

**DOI:** 10.1101/2021.03.23.21254159

**Authors:** Hana Mamo, Abinet Dagnaw, Nigussie Tadesse, Kalayu Berhane, Kehabtimer Shiferaw

**Affiliations:** Department of Public Health, College of Medicine and Health Sciences, Wachemo University, P.O.Box 667, Ethiopia; Department of Public Health, College of Health Science, Debre Berhan University, P.O.Box 77, Ethiopia; School of Public Health, Curtin University, Perth, Western Australia, Australia; School of Medicine, College of Health Sciences and Medicine, Wolaita Sodo University, P.O.Box 138, Ethiopia

**Keywords:** short inter-pregnancy interval, women, cross sectional, associated factors, Ethiopia

## Abstract

**Background:** Short inter-pregnancy interval is when the time elapsed between the dates of birth of the preceding child and the conception date of the current pregnancy is < 24 months. Despite its direct effects on the perinatal and maternal outcomes, there is a paucity of evidence on its prevalence and determinant factors, particularly in Ethiopia. Therefore, this study assessed the prevalence and associated factors of short inter-pregnancy interval among pregnant women in Debre Berhan town, Northern Ethiopia.

**Methods:** A community based cross sectional study was conducted among a randomly selected 496 pregnant women in Debre Berhan town from February 9 to March 9, 2020. The data was collected by using interviewer administered questionnaire and analyzed using STATA (14.2) statistical software. To identify the predictors of short inter-pregnancy interval, multivariable binary logistic regression was fitted and findings are presented using adjusted odds ratio (AOR) with 95% confidence interval (CI).

**Result:** The overall prevalence of short inter-pregnancy interval (<24 months) among pregnant women was 205 (40.9%). Being over 30 years of age at first birth (AOR=3.50; 95% CI: 2.12-6.01), non-use of modern contraceptive (AOR=2.51; 95% CI: 1.23-3.71), duration of breast feeding for less than 12 months (AOR= 2.62; 95% CI: 1.32-5.23), parity above four (AOR=0.31; 95% CI: 0.05-0.81), and unintended pregnancy (AOR= 5.42; 95% CI: 3.34-9.22) were independently associated factors with short inter-pregnancy interval.

**Conclusion:** Despite the family planning and other public health intervention tried in the country, the prevalence of short inter-pregnancy interval in this study was high. Therefore, it implies that increasing contraceptive use and encouraging optimal breast feeding might help in the efforts made to avert the problem.

## Introduction

The inter-pregnancy interval is the time between the birth of the preceding child and the conception date of the current pregnancy. A short inter-pregnancy interval is when the interval between the delivery date of the preceding live birth and the conception of date of the index birth is less than 24 months [1].

Historically, the World Health Organization (WHO) and other international authorities had recommended at least 2 to 3 years between successive pregnancies, and the United States Agency for International Development (USAID) had suggested an interval of 3 to 5 years. Given the inconsistency, various countries and regional programs requested the WHO to further review the research and provide recommendations. As a result, the report from the 2005 WHO Technical Consultation and Scientific Review of Birth Spacing recommended waiting at least 2 years after a live birth and 6 months after miscarriage or induced termination before conception of another pregnancy [1].

A short inter-pregnancy intervals (IPIs) is associated with adverse maternal and infant health outcomes. Short birth to pregnancy interval is known to hurt perinatal, neonatal and child health outcomes including: preterm birth, low birth weight, perinatal death, still birth, intellectual disability and developmental delay. Besides, it has also maternal health outcomes such as: nutritional depletion, anemia, cervical insufficiency, antepartum hemorrhage, premature rupture of membrane, and eclampsia [2-6].

In low-income countries, the prevalence of short inter-pregnancy interval ranges from 19.4% to 65.9% [4, 7]. Even though there are studies in developed and some low-income countries, there is paucity of evidence on the prevalence and predictors of inter-pregnancy interval in Ethiopia. On top of this, there are limitations to the currently available literature.

Most of the researches used birth interval, a proxy measure of time between two consecutive births, which could under or overestimate the time interval between the birth date of the preceding child and the conception date of the pregnancy. However, this study used the inter-pregnancy interval which measures correctly the time elapsed between the date of birth of the preceding child and the conception date of the current pregnancy. Therefore, the result of this study could present a true picture of the problem and aid in the efforts being tried to reduce the short inter-pregnancy interval. The aim of this study was to assess the prevalence and associated factors of short inter-pregnancy interval among pregnant women in Debre Berhan town, Northern Ethiopia.

## Methods and Materials

### Study setting, design and period

This community based cross-sectional study was conducted in Debre Berhan town, North Shewa zone, Northern Ethiopia from February 9 to March 9, 2020. The town is located 130 km from Addis Ababa, and it has fourteen kebeles.

### Populations

Pregnant women who gave at least one birth (uniparous) and live in Debre Berhan town were the source population of this study. Women who had miscarriage/abortion immediately before the current pregnancy were excluded from the study.

### Sample size determination and sampling procedure

The sample size was determined using Epi info version 7 considering 28.5 % of prevalence of short inter-pregnancy interval from the study done in Bahir Dar Felegehiwot Hospital [8]. The final sample size was determined as 517 after considering a design effect of 1.5 and a non-response rate of 10%. Simple random sampling technique was used to select the kebeles and the participants. Among the nine kebeles, five kebeles were randomly selected. Then the sample size was proportionally allocated to those five kebeles. Family folder from the hands of Health Extension Workers (HEWs) was used as a sampling frame to obtain a list of pregnant women in each kebele, and computer-generated random numbers were used to select the study participants.

### Data collection procedure and quality management

A structured interviewer administered questionnaire was prepared and implemented after reviewing relevant literatures. The questionnaire was prepared in English then translated to local language (Amharic) and finally translated back to English to check its consistency. It consists of socio-demographic, reproductive and health service related factors. It was checked and pre-tested in 5% of the study population outside the selected kebeles. After training was given for two days, the data was collected by five diploma midwives. Revisits of two to three times were made for women who were not available at the time of the survey. The collected data were checked for completeness and consistency on each days of data collection. Supervision and monitoring were made every day by the assigned supervisors and the principal investigator.

### Measurement

Inter-pregnancy interval was defined as the time in completed months from the reported date of live birth of the previous child to the self-reported last normal menstrual period (LNMP). Most participants knew the date of birth of the previous child and last normal menstrual period of the current pregnancy. However, in case of the participants who didn’t know the specific date of conception and/or the birth date of the previous child, the mid-date of the month was taken as the birth date of the previous child or date of the conception for the current pregnancy. Therefore, inter-pregnancy interval was calculated by subtracting the date of birth of the last child (previous child) from the date of conception of the current pregnancy (IPI= date of conception (LMP) - date of birth of the previous child). So, short inter-pregnancy interval was defined as an interval less than 24 months.

## Data processing and analysis

The data was entered, cleaned and processed by Epi-data version 3.1 software and exported to STATA version 14.2 for analysis. Descriptive statistics such as frequencies, proportions and summary statistics were used to describe the study population with relevant variables. Association between the outcome and explanatory variables was assessed by using a binary logistic regression model. Variables with p value of ≤ 0.2 in bivariable analysis were entered together to the model to conduct a multivariable analysis so as to control their effects of confounding. Statistical significance was considered at a level of significance of 5%, and adjusted odds ratio along with a 95% confidence interval was used to present the estimates of the strength of the associations. Hosmer-Lemeshow and variance inflation factor (VIF) was used to test the model fitness and multicollinearity respectively.

## Ethical consideration

Ethical clearance was obtained from the Ethical Review Committee of the Debre Berhan University. Then permission letter from the Debre Berhan town health office and Debre Berhan town administration office was obtained. Moreover, the informed written consent was obtained from each respondent. Personal identifier such as name was not mentioned in the questionnaire.

## Results

### Socio-demographic characteristics of the study participants

A total of 496 pregnant women were included in the study yielding a response rate of 96%. The age of the participants was ranged from 20 to 42 with the mean age (±SD) of 29.5 (±4.7) years. Of the study participants, around half (52.22%) were between the age of 25 to 29 years. One hundred seventy-three (34.88%) of the participants had attended college and above in their educational status, and 413(83.26%) were orthodox Christians by religion **(Table 1)**.

**Table 1:** Socio-demographic characteristics of the study participants, Debre Berhan town, Amahra region, Ethiopia, 2020.

| Characteristics | Frequency (n=496) | Percent (%) |
| --- | --- | --- |
| **Current maternal age** |  |  |
| 20-24 years | 43 | 8.66 |
| 25-29years | 259 | 52.22 |
| 30-34years | 110 | 22.17 |
| 35-39years | 61 | 12.29 |
| $\geq 40$years | 23 | 4.63 |
| **Maternal age at first birth** |  |  |
| ≤ 30 years | 415 | 83.67 |
| >30 years | 81 | 16.33 |
| **Religion** |  |  |
| Orthodox | 413 | 83.26 |
| Muslim | 39 | 7.86 |
| Protestant | 44 | 8.87 |
| **Current maternal occupational status** |  |  |
| Self employed | 81 | 16.33 |
| Private employee | 76 | 15.32 |
| Government employee | 147 | 29.64 |
| Housewife | 183 | 36.89 |
| Student | 9 | 1.81 |
| **Husband occupation** |  |  |
| Self employed | 155 | 31.25 |
| Private employee | 86 | 17.34 |
| Government employee | 236 | 47.58 |
| Other specify | 19 | 3.83 |
| **Household monthly income** |  |  |
| < 1000 ETB | 27 | 5.44 |
| 1000-2999 | 46 | 9.27 |
| 3000-4999 | 95 | 19.15 |
| > 5000 | 328 | 66.13 |

### Reproductive and health service related factors of the study participants

One hundred ninety-six (39.52%) of the participants did not use modern contraceptive before the current pregnancy, and 108 (21.77%) participants had an unintended pregnancy. Forty-eight (9.67%) of the study participants had no antenatal care (ANC) follow up by skilled attendants during the pregnancy of the index child. Similarly, forty-nine (9.87 %) of the participants provided breast feeding for their index child for only less than 12 months. Seventy-one (14.31%) participants had children of four and above excluding the current pregnancy **(Table 2)**.

**Table 2:**
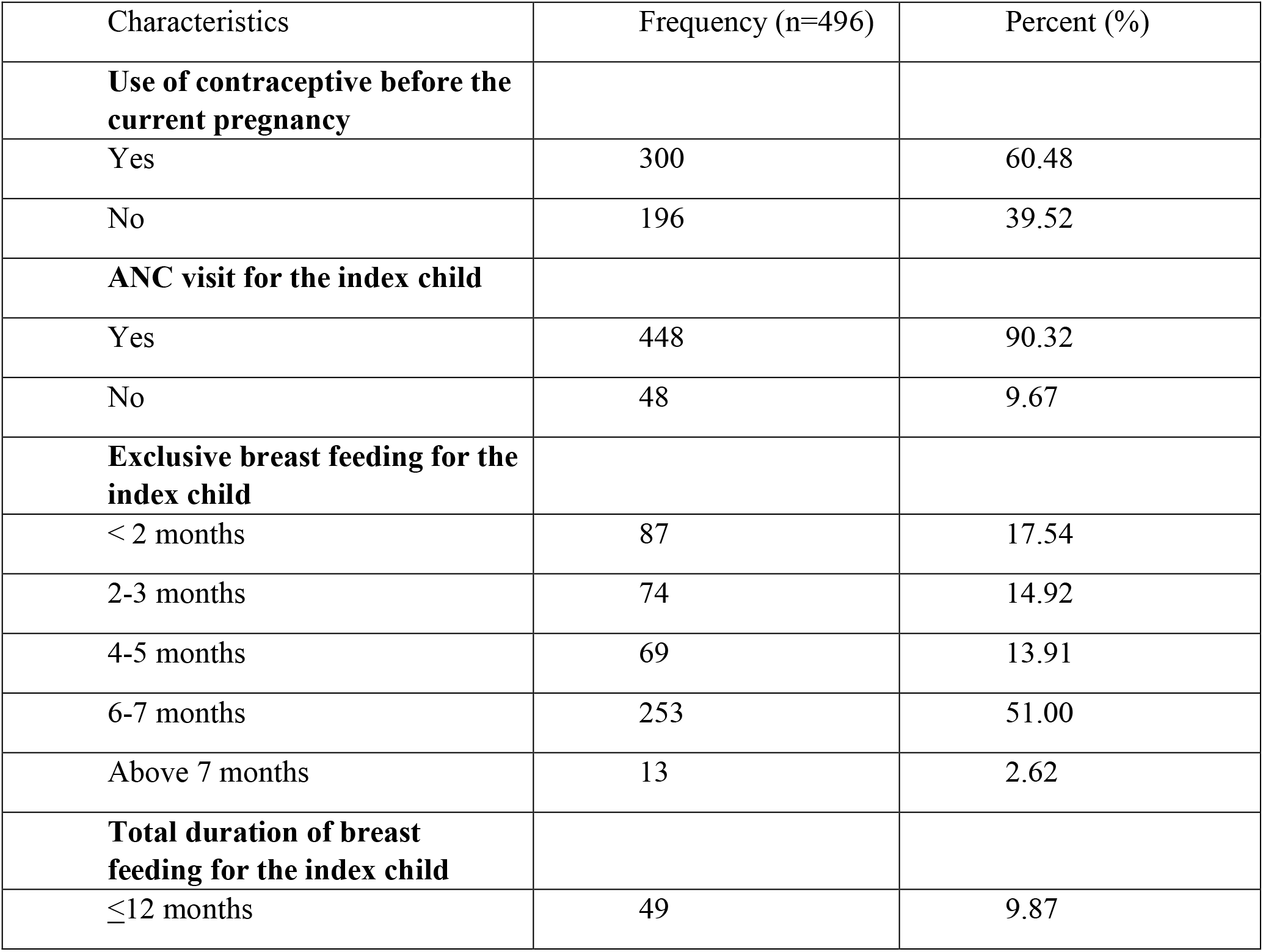
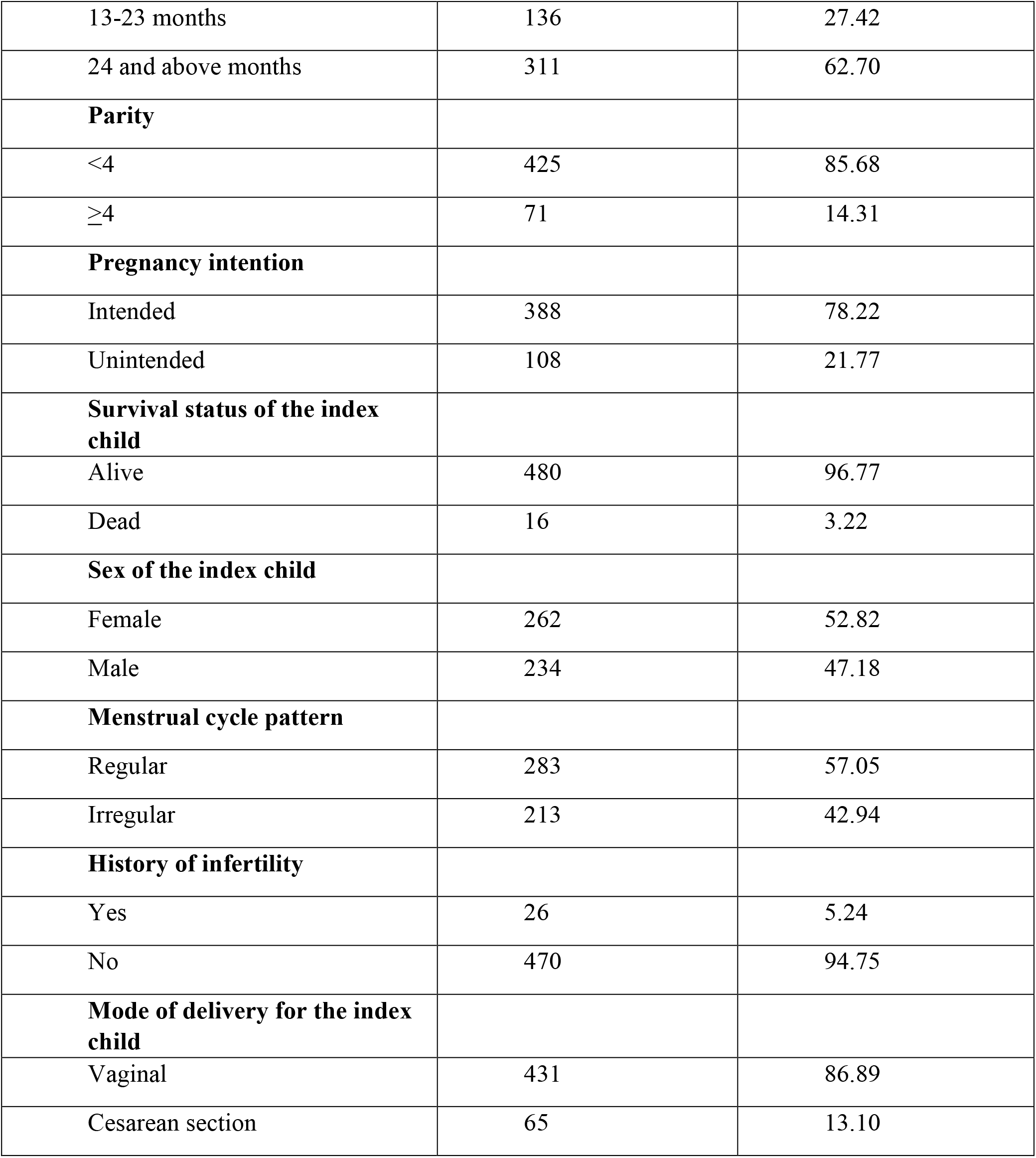
Reproductive and health service related factors of pregnant women in Debre Berhan town, Amahra region, Ethiopia, 2020

### Prevalence of short inter-pregnancy interval

The prevalence of short inter-pregnancy interval (< 24 months) of this study was 205 (40.9%) with 95% CI: 36.6 to 45.4%. The median inter pregnancy interval of the study participants was 29 (± IQR of 30) months. Of those who had short inter pregnancy interval, 24 (5%) had very short inter-pregnancy interval (<12 months). Besides, 210 (42%) and 86 (17.2%) of the participants had an inter-pregnancy interval of 24 to 59 months and more than 60 months, respectively.

### Factors associated with short inter-pregnancy interval

Bivariable and multivariable logistic regression analyses were carried out to determine the association between the explanatory variables and short inter-pregnancy interval. Hence, based on the p-value (< 0.2) of the bivariable analysis, current maternal age, age at first birth, parity, unintended pregnancy, non-use of modern contraceptive before the current pregnancy, duration of breast feeding, survival status of the index child were selected as candidate variables to be included in the final model. However, the result of multivariable analysis confirmed that age at first birth, parity, unintended pregnancy, non-use of modern contraceptive before the current pregnancy, and duration of breast feeding were independently associated with short inter-pregnancy interval. Multicollinearity was checked using a variance inflation factor and yielded a result of <10 for all variables in the final model. (Table 3)

**Table 3:**
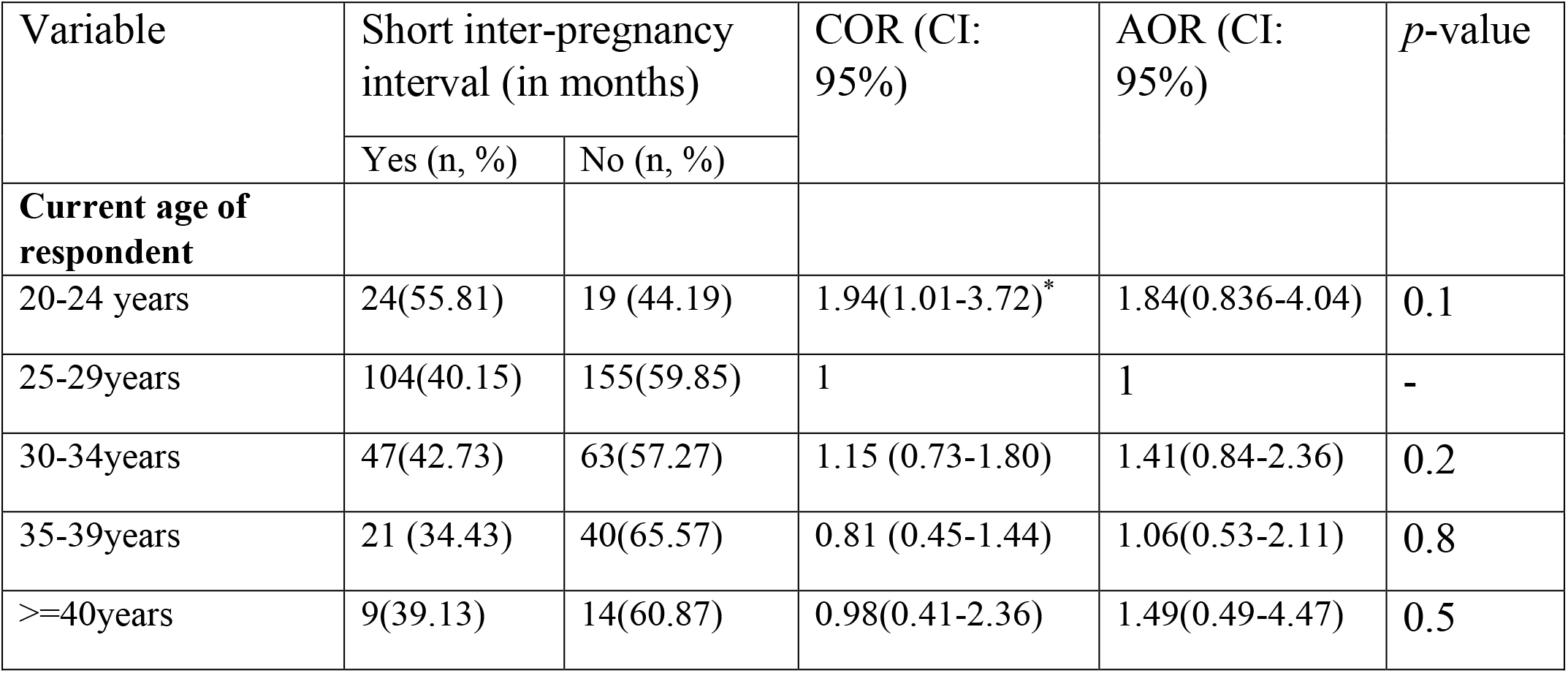
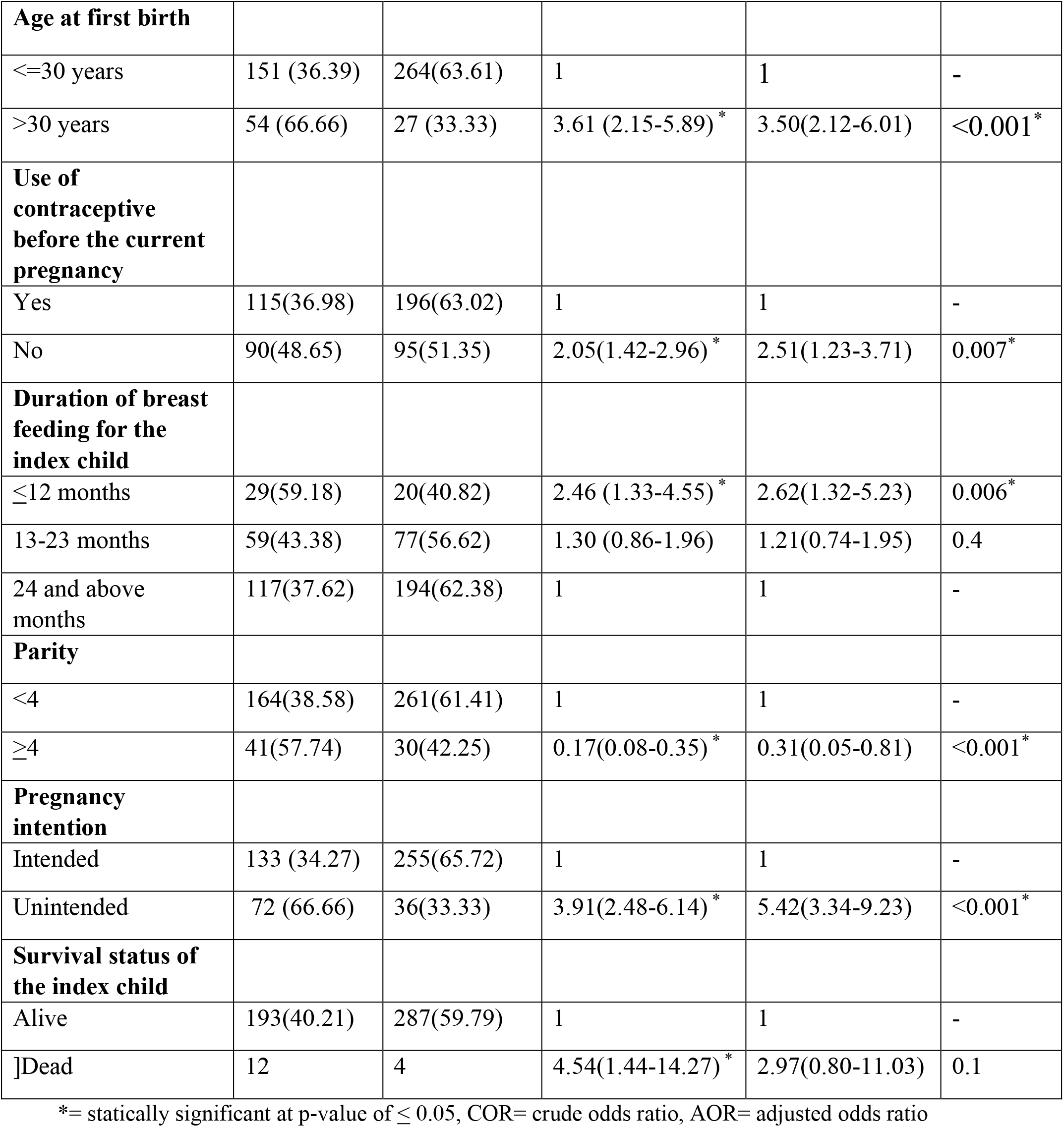
Bivariable and multivariable binary logistic regression analyses results of factors associated with short inter pregnancy interval among pregnant women in Debre Berhan town, Amahra region, Ethiopia, 2020

## Discussion

A community based cross-sectional study was conducted to assess the prevalence and associated factors of the short inter-pregnancy interval among pregnant women of Debre Berhan town.

Consequently, the overall prevalence of short inter-pregnancy interval (< 24 months) among pregnant women was 205 (40.9%). The factors independently associated with short inter-pregnancy interval were age at first birth, parity, unintended pregnancy, non -use of modern contraceptive before the current pregnancy, and duration of breastfeeding. The prevalence in this study is higher than the studies in Bahidar, Felegehiwot Hospital [8] and US[9] where about 28.5% and 35% of women had short inter-pregnancy interval respectively. This difference might be attributed to the cut off point for short inter-pregnancy interval. In this study, the cut off point for short inter-pregnancy interval was < 24 months. In comparison, the study conducted in US and RHC Manga Mandi, District Lahore defined short inter-pregnancy interval to be less than 18 months [9, 10]. On the other hand, this finding is lower compared to the study conducted in Nigeria [7] and Selangor [11] where the prevalence of short inter-pregnancy interval is 65.9% and 48% respectively. This difference might be attributed to the sample population and socio-cultural practice.

In this study, the odds of experiencing short inter-pregnancy interval was 3.5 times higher among women who started child bearing above the age of 30 years compared to those who start at 30 years of age and lower. This finding is consistent with the study done in Bahirdar (Felegehiwot hospital) [8], US [9] and Michigan [12]. This might be due to the intention to use the remaining fertility age efficiently before the woman reaches the stage of menopause. In line with the evidence from two studies done in Port Harcourt [13] and Nigeria [7], the finding of this study revealed that women who did not use modern contraceptive before the current pregnancy had 2.5 times higher odds to experience short inter-pregnancy interval as compared to those who used it. This might be due to a woman who used any type of the modern contraceptive has a probability of increasing the interval between the previous birth and the next pregnancy.

This study also found out unintended pregnancy to be associated with short inter-pregnancy interval. The odds of experiencing short inter-pregnancy interval was 5.4 times higher among women with unintended current pregnancy compared to their counterparts. This finding is congruent with the study conducted in the US [9] and Selangor [11]. This might be due to a woman who plan to be pregnant may follow the recommendation for child spacing and therefore end up with optimal inter-pregnancy interval. “Since non-utilization and failure of contraceptive are among the major contributors of the unintended pregnancy, this might have contributed to the shortened inter-pregnancy interval.” [14]. This study revealed that the odds of short inter-pregnancy interval was 2.6 times higher among women who breast fed their last child for less than 12 months compared to those who breast fed for 24 and above months. This finding is in line with the evidence from the study done in Nigeria [7]. It might be due to the fact that duration of breast feeding including exclusive breast feeding improves infant survival and lengthens the interval between pregnancies due to lactational amenorrhea (negative hormonal feedback). During breastfeeding, the receptors in the breast nipple will be stimulated, and this initiates a signal to the hypothalamus: a nerve center in the brain which in turn signals the pituitary gland thereby inhibits ovulation by reducing the release of gonadotrophic hormone needed for ovulation which results in post-partum amenorrhea [15].

The study also showed that parity was associated with short inter-pregnancy interval. Women who had four and above children had 70% lower odds to experience short inter pregnancy interval compared to the counter groups. This finding is in line with the study done in Nigeria [13, 16] and rural Bangladesh[16], but in contrast with the study done in Selangor [11]. These women may have achieved their desired family size and may feel less pressure or may be in less hurry to get pregnant again.

## Limitation

The inter-pregnancy interval and breast-feeding duration were calculated based on women recall, which might result in recall bias. Being the data obtained through self-report of the women, the accuracy might not be as a level obtained objectively, even though respondents were critically informed about giving accurate information through assuring the confidentiality of their responses. The exclusion of women who experienced miscarriage/abortion immediately before the current pregnancy might have underestimated the prevalence of the short inter pregnancy interval.

## Conclusion

The World Health Organization (WHO) and the government of Ethiopia recommended that a woman should wait 24 months before attempting the next pregnancy after a live birth. Despite this recommendation, this study found out a higher proportion of women (40.9%) getting pregnant before the recommended period of time. Age at the first birth, parity, non -use of modern contraceptive, duration of breast feeding, and unintended pregnancy were independently associated with short inter-pregnancy interval in the study. Therefore, it implies that increasing contraceptive use and encouraging optimal breast feeding might help in the efforts made to avert the problem. Besides, further studies in the rural setup with higher sample size are needed to ascertain the prevalence and determinants of short inter-pregnancy interval.

## Data Availability

All the supplementary data can be obtained from the corresponding author up on request.

